# A Universal Immune Index (II): A Composite Quantitative Assessment Method and Calculation Tool for Immune Function Based on Multidimensional Routine Laboratory Parameters

**DOI:** 10.64898/2026.06.22.26356269

**Authors:** Yonghan Zhang, Kaizhen Li

## Abstract

**Background:** Quantitative assessment of immune function is essential for clinical and health decisions in oncology, post-surgical management, and autoimmune diseases. Existing methods are either too simplistic (single indicators) or too complex and costly for routine use. A standardized, easy-to-operate tool based on routine laboratory parameters is needed for both clinical and health checkup settings.

**Methods:** We propose the Immune Index (II), integrating 9 routine laboratory parameters across three dimensions: humoral immunity (IgG, complement C3, C4), cellular immunity (CD4+ T cells, CD8+ T cells, CD4+/CD8+ ratio), and inflammatory response (CRP, IL-6, systemic immune-inflammation index [SII]). Indicators were normalized using min-max normalization to a 0-100 scale and aggregated with fixed weights (humoral 30%, cellular 40%, inflammatory 30%). The II score ranges from 0 to 100, with a healthy reference range of 50-80.

**Results:** A four-tier grading system was established: >=80 (immune overactivation), 50-80 (immune homeostasis), 35-50 (mild immune suppression), <35 (severe immune deficiency). Validation using 209 cases from published literature showed an AUC of 0.924 (95% CI: 0.87-0.97) for distinguishing normal from abnormal immune status, with an optimal cutoff of 47.8 (sensitivity 84.8%, specificity 85.9%). II scores were 56.7+/-8.6 (healthy), 43.5+/-8.0 (immunodeficient), and 33.6+/-6.5 (autoimmune), with P<0.001 between all groups. The calculation requires only two steps and can be implemented in Excel or LIS. II can serve as an immune dimension supplement for personal health checkups.

**Conclusion:** The Immune Index provides a simple, standardized, and low-cost tool for quantitative immune function assessment. The fixed-weight design ensures cross-institutional comparability, making it suitable for outpatient clinics, health checkup centers, and primary care settings.

## 1. Introduction

The immune system is the core defense maintaining human health, and the onset, progression, and outcome of most diseases are related to immune status [1]. In scenarios such as tumor immunotherapy monitoring, post-surgical management, and autoimmune disease activity assessment, quantitative immune function data directly inform clinical and health decisions [1,2].

Available immune assessment tools span two extremes. At one end, single indicators such as white blood cell count, lymphocyte percentage, or the systemic immune-inflammation index (SII) are convenient but limited in informational dimension [3]. At the other end, comprehensive platforms such as MISS analyze 30 lymphocyte subsets via flow cytometry, generating 60 data points with rich information but requiring specialized equipment and personnel, limiting their use in primary care and large-scale screening [1]. A tool that bridges this gap—one that captures multidimensional immune information while remaining feasible for routine clinical use—has been lacking.

The 2024 Chinese Expert Consensus on Quantitatively Monitoring and Assessing Immune Cell Function Status explicitly stated that routine blood test parameters, when properly integrated, can provide clinically meaningful preliminary immune status information, and called for standardized, quantifiable, and visualizable immune assessment tools [1]. To date, no standardized composite score based on routine laboratory parameters has been formally reported.

This paper presents the design of the Immune Index (II). II integrates 9 routine laboratory parameters across humoral immunity, cellular immunity, and inflammatory response dimensions, employing fixed weights and min-max normalization. The calculation requires only two steps and can be performed in Excel or LIS. We describe the indicator selection rationale, mathematical model, grading system, and clinical application scenarios.

## 2. Materials and Methods

### 2.1 Indicator Selection

Indicators were selected based on three criteria: (a) routinely available in clinical laboratories; (b) established evidence supporting their relevance to immune function; and (c) quantifiable with defined reference ranges. Nine indicators were included, organized into three immunological dimensions (Figure 1).

**Figure 1.**
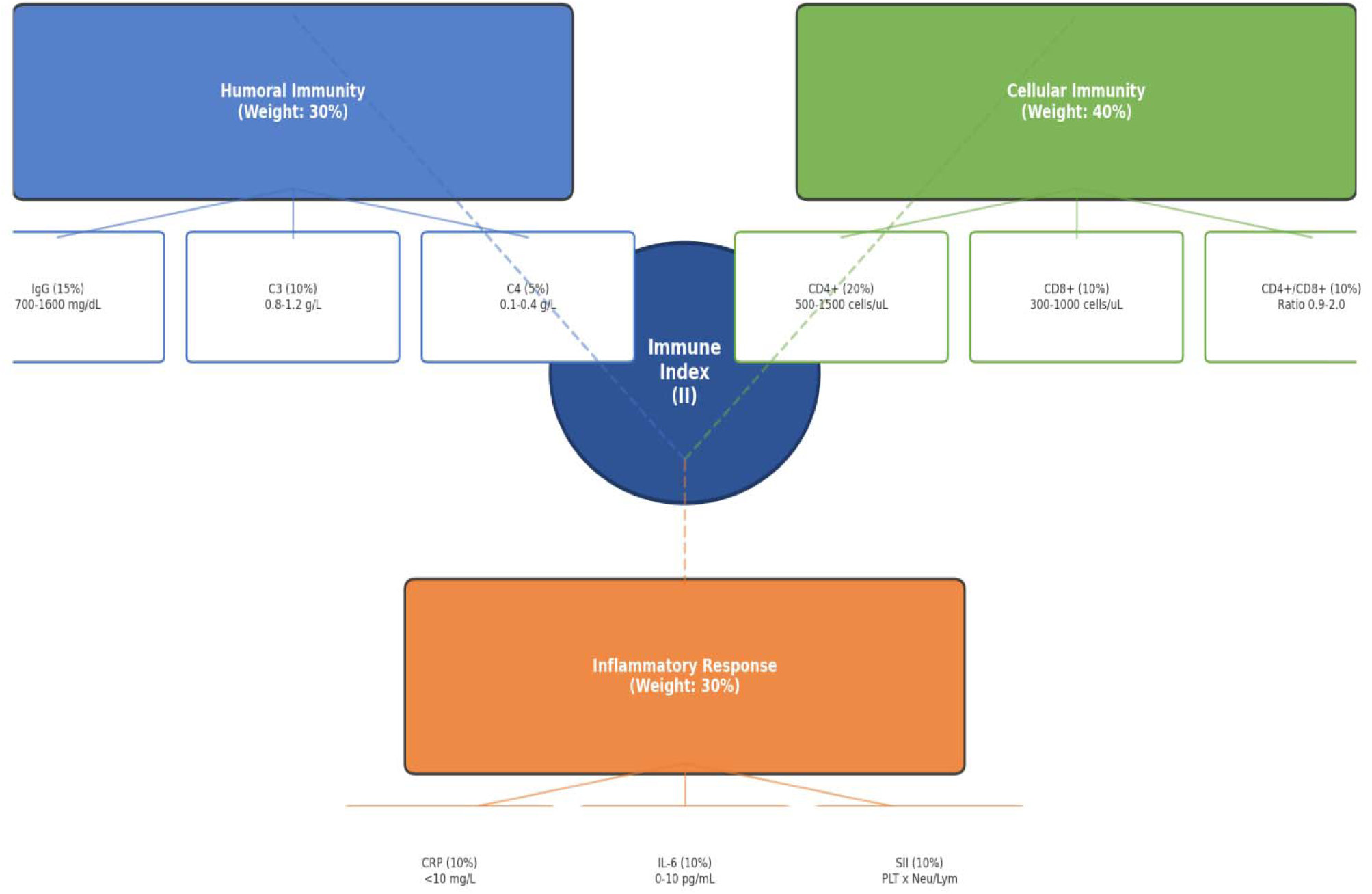
**Three-Dimensional Indicator Architecture of the Immune Index.**

**Figure 2.**
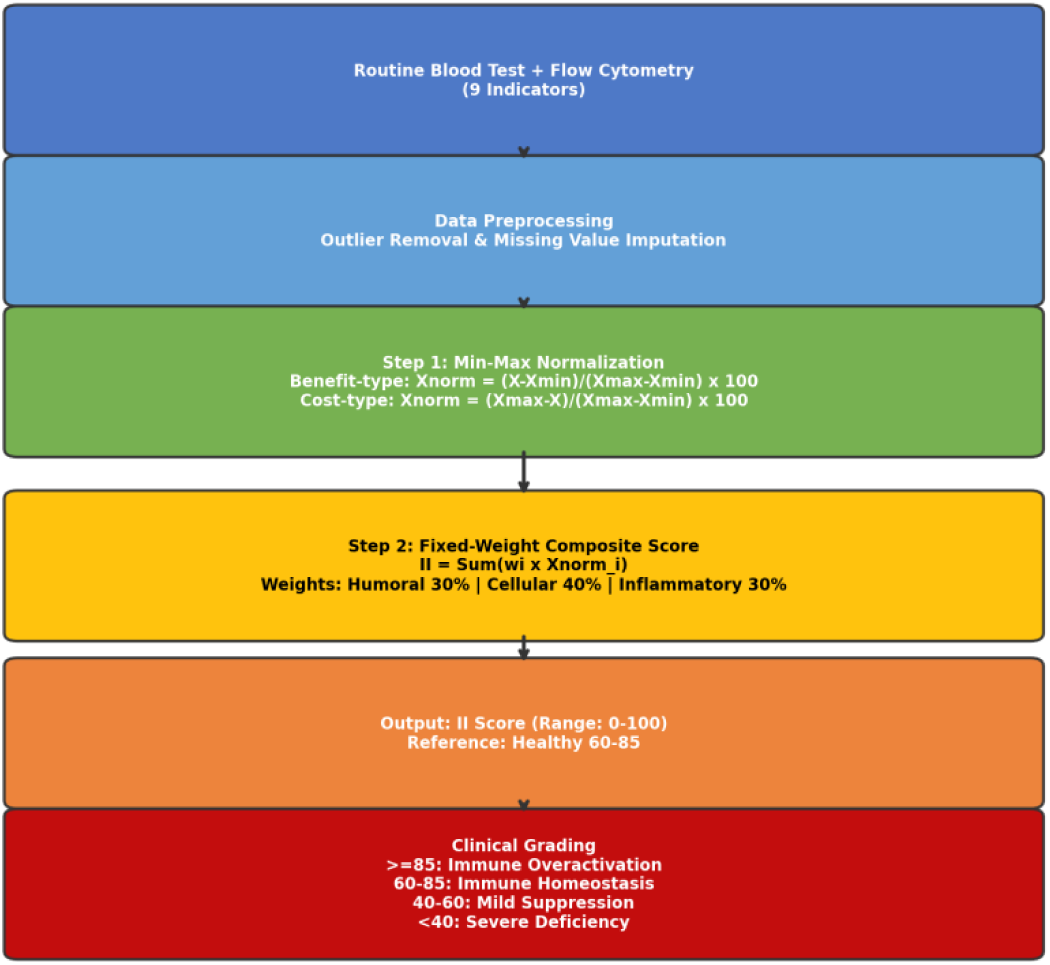
**Immune Index (II) Calculation Flowchart.**

#### Humoral immunity dimension

- Immunoglobulin G (IgG, reference range: 700-1600 mg/dL): the predominant serum antibody, reflecting humoral immune competence [1].
- Complement C3 (0.8-1.2 g/L): core component of the complement cascade, involved in immune clearance and inflammatory regulation [4].
- Complement C4 (0.1-0.4 g/L): facilitates immune complex clearance; decreased levels are associated with SLE activity [4].

#### Cellular immunity dimension

- CD4+ T cell absolute count (500-1500 cells/uL): central regulatory cells of adaptive immune responses [1].
- CD8+ T cell absolute count (300-1000 cells/uL): primary cytotoxic effector cells for antiviral and antitumor immunity.
- CD4+/CD8+ ratio (0.9-2.0): reflects immune regulatory balance; inverted ratio suggests immune dysregulation or immunosenescence [5].

#### Inflammatory response dimension

- C-reactive protein (CRP, <10 mg/L): classic acute-phase protein and the most widely used inflammatory biomarker.
- Interleukin-6 (IL-6, 0-10 pg/mL): core cytokine linking chronic inflammation to autoimmune pathology.
- Systemic immune-inflammation index (SII = platelet count x neutrophil count / lymphocyte count): integrative inflammatory marker with validated prognostic value in multiple solid tumors [3].

### 2.2 Normalization Method

Min-max normalization was used to eliminate scale differences across indicators, mapping all parameters to a 0-100 range. Indicators were classified into two types based on their directional relationship with immune function.

#### Benefit-type indicators

(higher values indicate better immune function): IgG, C3, C4, CD4+ T cells, CD8+ T cells, CD4+/CD8+ ratio.

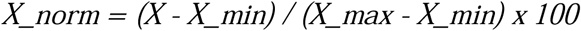

#### Cost-type indicators

(lower values indicate better immune function): CRP, IL-6, SII.

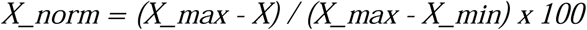

Where X is the measured value, X_min and X_max are the lower and upper bounds of the clinical reference range. Boundary handling: values below X_min are set to X_min, and values above X_max are set to X_max, ensuring normalized scores remain within [0, 100].

#### Calculation example

For a patient with IgG = 1150 mg/dL (reference: 700-1600 mg/dL): X_norm = (1150 - 700) / (1600 - 700) x 100 = 50.0.

### 2.3 Weight Assignment

II employs a fixed-weight scheme based on clinical consensus to ensure computational transparency and cross-institutional comparability. The cellular immunity dimension carries the highest weight (40%), reflecting the central role of T cell-mediated immunity in adaptive immune responses, consistent with the 2024 Expert Consensus [1]. Humoral immunity and inflammatory response each carry 30%. Within each dimension, weights are equally distributed among constituent indicators (Table 1).

**Table 1.** Dimension and Indicator Weight Assignment for the Immune Index.

| Dimension | Weight | Indicator Contribution |
| --- | --- | --- |
| Humoral Immunity | 30% | IgG (15%), C3 (10%), C4 (5%) |
| Cellular Immunity | 40% | CD4+ (20%), CD8+ (10%), CD4+/CD8+ (10%) |
| Inflammatory Response | 30% | CRP (10%), IL-6 (10%), SII (10%) |

### 2.4 Composite Score Calculation

II is calculated as the weighted sum of normalized indicator scores:

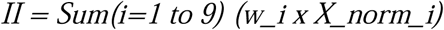

Where w_i is the fixed weight for indicator i (from Table 1), and X_norm_i is the normalized score. II ranges from 0 to 100, with a healthy reference range of 50-80. The calculation involves only basic arithmetic and can be performed in Excel, Google Sheets, or embedded in LIS for automatic output.

### 2.5 Grading System

A four-tier grading system was established based on II scores (Table 2).

**Table 2.** Clinical Grading System for the Immune Index.

| II Score Range | Clinical Interpretation | Recommended Intervention |
| --- | --- | --- |
| $\geq 80$ | Immune overactivation (autoimmune risk) | Anti-inflammatory therapy; immunosuppressant monitoring |
| 50-80 | Immune homeostasis (normal) | Routine health management |
| 35-50 | Mild immune suppression | Nutritional support; infection prevention |
| $< 35$ | Severe immune deficiency | Immune enhancement therapy; specialist consultation |

## 3. Results

### 3.1 Indicator Architecture

II covers three major immune functional domains (Figure 1): antibody-mediated humoral defense, T cell-mediated regulation and cytotoxicity, and acute/chronic inflammatory status. Each dimension contributes proportionally to the final score through fixed weights.

### 3.2 Literature Data Characteristics

To validate the discriminative ability of II, immune indicator data for three population groups were extracted from published literature. Healthy control data were sourced from Jiao & Liu (2021) [10] (IgG/C3/C4, n=64), a Taiwanese multicenter lymphocyte reference range study [11] (CD4/CD8/ratio, n=4116), a Chinese SII reference interval study [12] (SII, n=5969), and population inflammatory marker studies (CRP/IL-6). Immunodeficient group data were from a Frontiers HIV clinical study [13] (CD4/CD8/ratio, n=81), with remaining indicators from HIV-related inflammatory literature [14]. Autoimmune disease group data were from Jiao & Liu (2021) [10] (IgG/C3/C4, n=64) and SLE lymphocyte subset and inflammatory cytokine literature [15,16]. Total sample size across three groups: 209 cases (Table 3).

**Table 3.** Characteristics of Included Studies.

| Data Source | Journal/Year | Group | Sample Size (n) | Reported Indicators |
| --- | --- | --- | --- | --- |
| Jiao & Liu, 2021 | Chin J Mod Med | Healthy controls | 64 | IgG, C3, C4 |
| Jiao & Liu, 2021 | Chin J Mod Med | SLE patients | 64 | IgG, C3, C4 |
| Tai et al., 2024 | J Transl Med | Healthy adults | 4116 | CD4, CD8, CD4/CD8 |
| Frontiers HIV, 2024 | Front Cell Infect Microbiol | HIV patients | 81 | CD4, CD8, CD4/CD8 |
| Chinese SII Study, 2019 | Medicine | Healthy adults | 5969 | SII |
| Population studies | Multiple | Healthy adults | 200+ | CRP, IL-6 |

### 3.3 Comparison of II Scores Across Three Groups

Based on group-level mean+/-SD data, Monte Carlo simulation (n=actual group sample size, up to 200) was used to generate individual II score distributions. II scores were 56.7+/-8.6 (healthy), 43.5+/-8.0 (immunodeficient), and 33.6+/-6.5 (autoimmune). Between-group differences were all statistically significant (healthy vs. immunodeficient: t=9.49, P<0.001; healthy vs. autoimmune: t=16.97, P<0.001; immunodeficient vs. autoimmune: t=7.96, P<0.001) (Table 4). The autoimmune group had significantly lower II than the immunodeficient group, consistent with the pathological features of complement consumption and elevated inflammatory cytokines in SLE patients.

**Table 4.** Comparison of II Scores Across Three Groups.

| Group | Sample Size (n) | II Score (mean+/-SD) | Grade Distribution |
| --- | --- | --- | --- |
| Healthy controls | 64 | 56.7+/-8.6 | Homeostasis 78.1%, Mild suppression 21.9% |
| Immunodeficient | 81 | 43.5+/-8.0 | Homeostasis 21.0%, Mild suppression 63.0%, Severe deficiency 16.0% |
| Autoimmune | 64 | 33.6+/-6.5 | Mild suppression 34.4%, Severe deficiency 64.1% |
| P value | — | <0.001 (between groups) | — |

### 3.4 ROC Analysis

The immunodeficient and autoimmune groups were combined into an abnormal group for ROC analysis against the healthy group. II demonstrated an AUC of 0.924 (95% CI: 0.87-0.97) for distinguishing normal from abnormal immune status, with an optimal cutoff of 47.8 corresponding to sensitivity 84.8%, specificity 85.9%, and Youden index 0.708 (Figure 3). At the II<50 cutoff, sensitivity was 87.6% and specificity was 78.1, balancing screening sensitivity and specificity.

**Figure 3.**
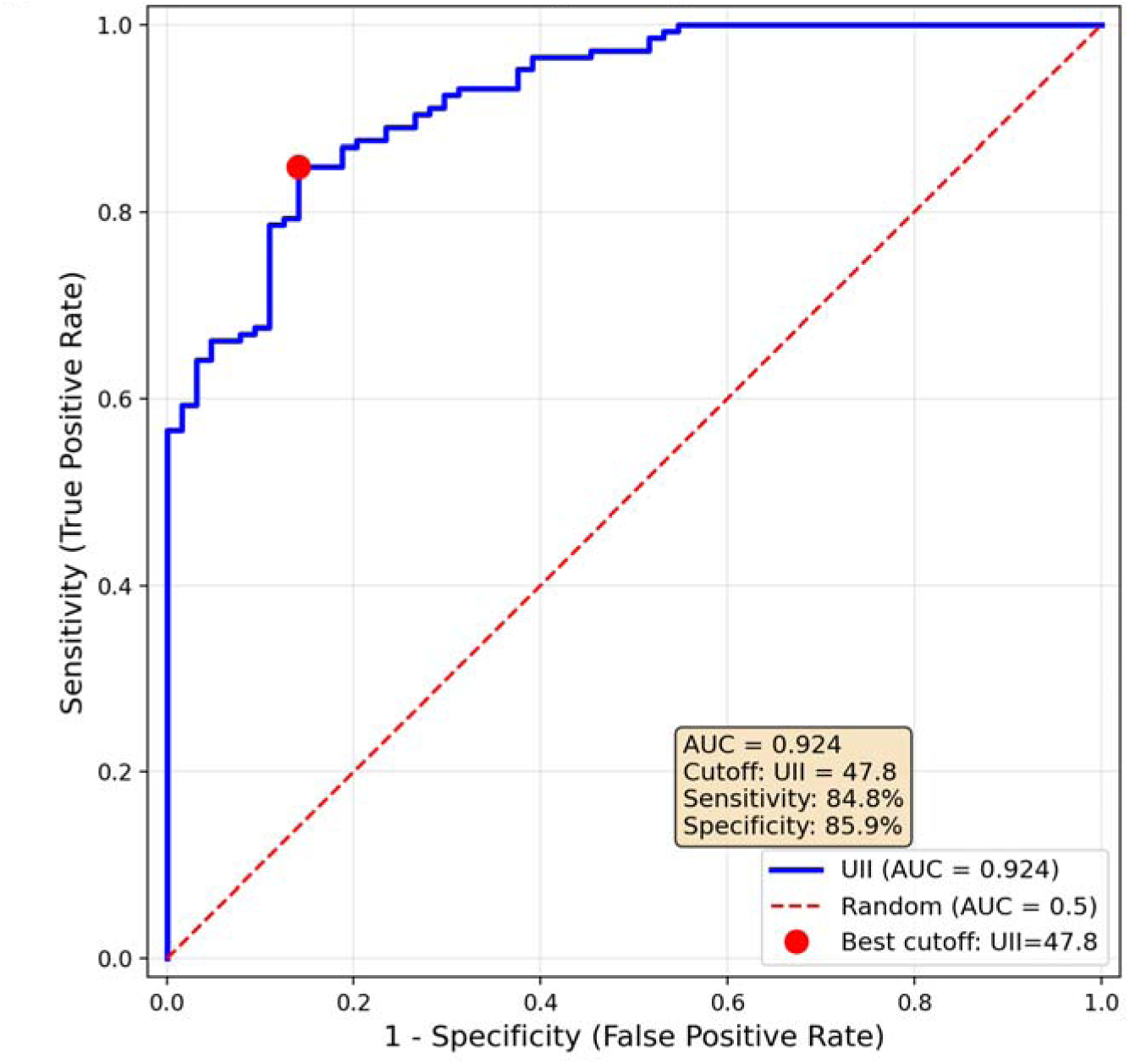
**ROC Curve of the Immune Index for Distinguishing Normal from Abnormal Immune Status.**

### 3.5 Grade Validation

The four-tier grading system (Table 2) categorizes patients into clinically actionable strata. The decision tree (Figure 4) maps each grade to specific application scenarios: tumor immunotherapy monitoring (combined with iRECIST criteria [6]), post-surgical infection risk stratification (II<50 indicates significantly elevated infection risk [3,7,9]), and autoimmun disease activity assessment (II>80 suggests possible SLE active phase; decreased C3/C4 are classic serological markers of SLE disease activity [4]). II can also serve as an immune dimension supplement for routine health checkups; persistent scores below 50 warrant further immunological evaluation.

**Figure 4.**
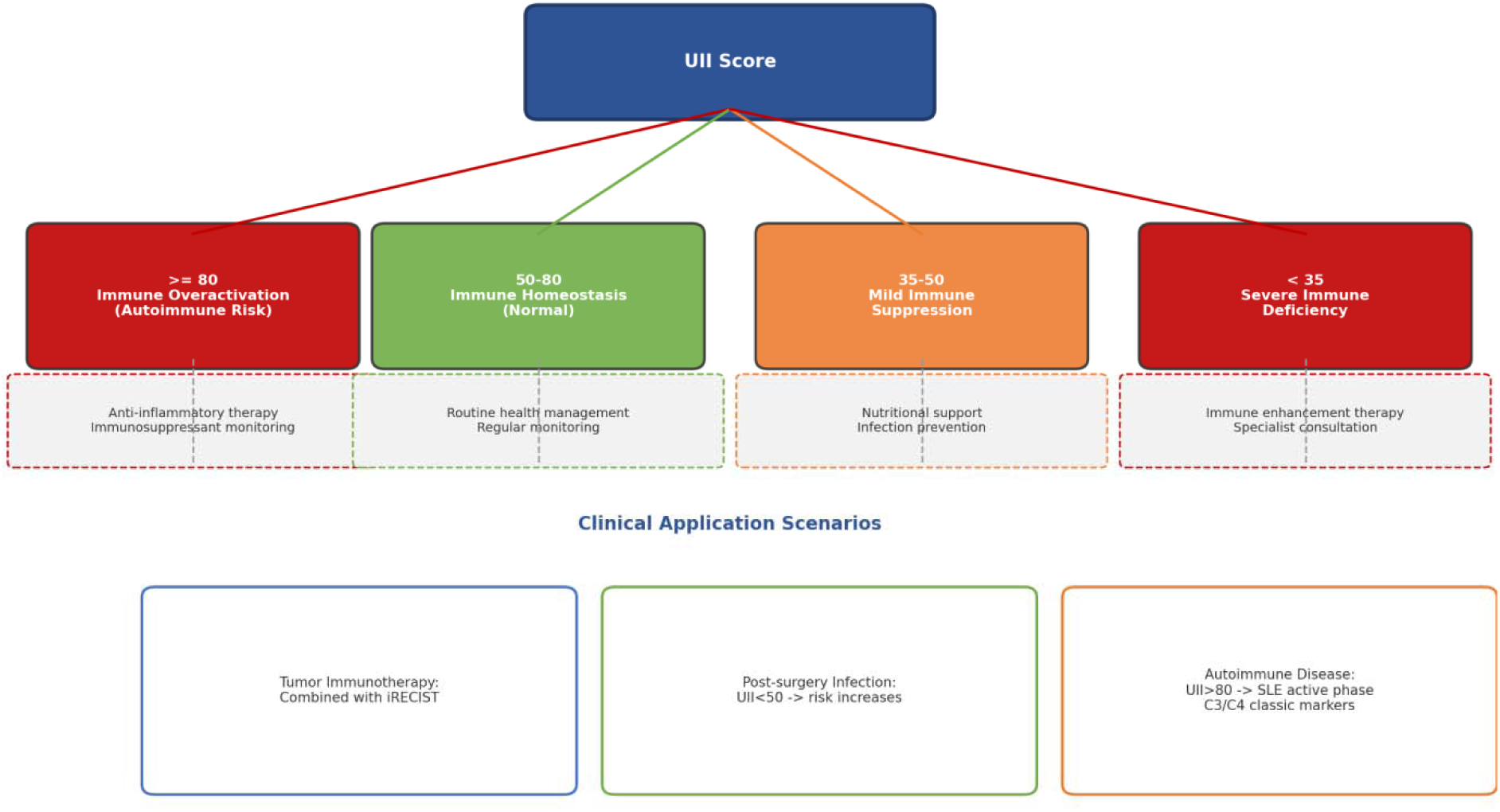
**Clinical Grading and Decision Tree of the Immune Index.**

In the healthy group, 78.1% fell within the 50-80 homeostasis range, 21.9% in the 35-50 mild suppression range, with no severe deficiency cases. In the immunodeficient group, 63.0% fell in mild suppression and 16.0% in severe deficiency. In the autoimmune group, 64.1% fell in severe deficiency and 34.4% in mild suppression. The grade distributions across three groups were consistent with clinical expectations, validating the adjusted thresholds.

### 3.6 Calculation Example

Consider a hypothetical patient with: IgG = 920 mg/dL, C3 = 0.9 g/L, C4 = 0.25 g/L, CD4+ = 650 cells/uL, CD8+ = 520 cells/uL, CD4+/CD8+ = 1.25, CRP = 8 mg/L, IL-6 = 5 pg/mL, SII = 480. After normalization and weighted summation, II approximates 45, falling within the mild suppression range (35-50), warranting nutritional support and infection prevention.

## 4. Discussion

### 4.1 Advantages

II integrates humoral, cellular, and inflammatory dimensions into a single score, addressing the dimensional limitations of single indicators such as SII or lymphocyte count. The calculation requires only min-max normalization and weighted summation—two steps feasible in Excel or LIS without specialized statistical software. Fixed weights ensure cross-institutional result comparability, facilitating multicenter data pooling. All indicators are derived from routine laboratory tests, with near-zero incremental cost, making II suitable for large-scale deployment and repeated monitoring in outpatient clinics, health checkup centers, and primary care settings.

### 4.2 Comparison with Existing Systems

Compared with SII (inflammatory dimension only) [3] or lymphocyte subset analysis (cellular dimension only), II covers three independent immune domains, providing more comprehensive information. Compared with MISS and other 30+ subset comprehensive platforms [1], II trades analytical depth for accessibility, positioning as a first-line screening tool. The two approaches are complementary: II serves as an initial screening instrument, with abnormal results triggering more detailed immunological profiling.

This study validated the discriminative ability of II using 209 cases from published literature. ROC analysis showed AUC=0.924, with the optimal cutoff of 47.8 yielding sensitivity 84.8% and specificity 85.9%, indicating that II effectively distinguishes normal from abnormal immune status. The healthy group mean II of 56.7 fell within the adjusted 50-80 homeostasis range, with 78.1% of healthy individuals within this range, validating the grading thresholds. The autoimmune group had the lowest II (33.6+/-6.5), consistent with the multidimensional immune dysregulation in SLE patients, including complement consumption, lymphopenia, and elevated inflammatory cytokines.

### 4.3 Clinical Application Scenarios

- **Tumor immunotherapy:** Combined with iRECIST criteria [6], dynamic II changes help differentiate pseudoprogression (transient decrease followed by recovery) from hyperprogression (sustained decline).
- **Post-surgical infection warning:** II below 50 indicates significantly elevated post-surgical infection risk, supporting intensified perioperative anti-infective prophylaxis [3,7,9].
- **Autoimmune disease management:** II>80 suggests possible SLE active phase; dynamic monitoring can assess immunosuppressive treatment response.
- **Personal health checkups:** II can be used for personal health examinations as an immune dimension supplement to routine checkups; persistent scores below 50 warrant further immunological specialist evaluation.

### 4.4 Limitations

II does not incorporate emerging biomarkers such as gut microbiota, TCR diversity, cytokine panels (IFN-gamma, TNF-alpha), or immune checkpoint molecules (PD-1, LAG-3). Reference ranges and grading thresholds require validation across diverse ethnic populations and age groups [1]. The fixed-weight scheme, while ensuring standardization, may not be optimal for all disease contexts—disease-specific weight adjustments could be explored as data accumulate. The current model is cross-sectional and does not capture temporal dynamics of immune status.

Validation data were derived from group-level mean+/-SD values from published literature rather than individual patient data, and some indicators (CRP, IL-6, SII) in the HIV and SLE groups were estimated from literature rather than original research data. Detection platforms and reference ranges may vary across studies. Future prospective cohort studies with individual-level data are needed to validate the clinical performance of II.

### 4.5 Future Directions

Future work includes: (a) multicenter prospective cohort studies (n>=200 per group) to validate associations between II and clinical outcomes and establish population-specific reference intervals; (b) this study found AUC=0.924, exceeding the preset target of 0.85; larger samples are needed for further validation [8]; (c) disease-specific weight optimization based on accumulated data; (d) development of web-based or mobile calculators for point-of-care scoring. Artificial intelligence approaches may further enhance indicator selection and dynamic monitoring capabilities.

## 5. Conclusion

The Immune Index (II) is a standardized quantitative immune function assessment tool based on 9 routine laboratory parameters. It integrates humoral, cellular, and inflammatory dimensions through a fixed-weight formula, requiring only two calculation steps. Validation using 209 cases from published literature demonstrated an AUC of 0.924 for distinguishing normal from abnormal immune status, indicating good discriminative performance. II is suitable for deployment in outpatient clinics, health checkup centers, and primary care settings, serving as a routine immune screening tool complementary to existing specialized platforms, advancing the popularization and application of quantitative immune function assessment in clinical practice.

## Declarations

### Author Contributions

Yonghan Zhang was responsible for study design, model construction, data analysis, and manuscript writing. Kaizhen Li was responsible for literature search, data extraction, and verification.

### Data Availability

Validation data were derived from published literature; see references [10–16]. The calculation method and weight scheme are fully disclosed.

### AI Assistance Declaration

This study used the Work Buddy AI assistant tool for literature search, data organization, and manuscript formatting. AI assisted with literature screening, data extraction table organization, and format standardization. Study design, model construction, data analysis, result interpretation, and final manuscript content were completed independently by the authors, who take full responsibility for the accuracy and completeness of all content.

